# Persons Diagnosed with COVID in England in the Clinical Practice Research Datalink (CPRD): A Cohort Description

**DOI:** 10.1101/2023.03.17.23287415

**Authors:** Kathleen M. Andersen, Leah J. McGrath, Maya Reimbaeva, Diana Mendes, Jennifer L. Nguyen, Kiran K. Rai, Theo Tritton, Carmen Tsang, Deepa Malhotra, Jingyan Yang

## Abstract

**Objective:** To create case definitions for confirmed COVID diagnoses, COVID vaccination status, and three separate definitions of high risk of severe COVID, as well as to assess whether the implementation of these definitions in a cohort reflected the sociodemographic and clinical characteristics of COVID epidemiology in England.

**Design:** Retrospective cohort study

**Setting:** Electronic healthcare records from primary care (Clinical Practice Research Datalink, or CPRD) linked to secondary care data (Hospital Episode Statistics, or HES) data covering 24% of the population in England

**Participants:** 2,271,072 persons aged 1 year and older diagnosed with COVID in CPRD Aurum between August 1, 2020 through January 31, 2022.

**Main Outcome Measures:** Age, sex, and regional distribution of COVID cases and COVID vaccine doses received prior to diagnosis were assessed separately for the cohorts of cases identified in primary care and those hospitalized for COVID (primary diagnosis code of ICD-10 U07.1 “COVID-19”). Smoking status, body mass index and Charlson Comorbidity Index were compared for the two cohorts, as well as for three separate definitions of high risk of severe disease used in the United Kingdom (NHS Highest Risk, PANORAMIC trial eligibility, UK Health Security Agency Clinical Risk prioritization for vaccination).

**Results:** Compared to national estimates, CPRD case estimates underrepresented older adults in both the primary care (age 65-84: 6% in CPRD vs 9% nationally) and hospitalized (31% vs 40%) cohorts, and overrepresented people living in regions with the highest median wealth areas of England (20% primary care and 20% hospital admitted cases in South East, vs 15% nationally). The majority of non-hospitalized cases and all hospitalized cases had not completed primary series vaccination. In primary care, persons meeting high risk definitions were older, more often smokers, overweight or obese, and had higher Charlson Comorbidity Index score.

**Conclusions:** CPRD primary care data is a robust real-world data source and can be used for some COVID research questions, however limitations of the data availability should be carefully considered. Included in this publication are supplemental files for atotal of over 28,000 codes to define each of three definitions of high risk of severe disease.

**SUMMARY BOXES:** *What is already known on this topic?:*

- The UK Government publishes data on cases, hospital admissions and vaccinations related to COVID in England.
- There are at least three definitions of persons at high-risk of severe COVID in use in England.

*What this study adds:*

- Our study created case definitions for COVID diagnoses, COVID vaccination, and three separate definitions of high risk of severe COVID for use in the Clinical Practice Research Datalink (CPRD), a database covering 24% of England.
- The COVID population in the CPRD has a different age and regional distribution than national case reports, which future studies may need to consider.

## INTRODUCTION

As of 3 February 2023, there have been over 20 million confirmed COVID cases and more than 183,000 related deaths in England.^1^ COVID severity ranges from asymptomatic cases to severe disease requiring hospital admission and sometimes death, with older adults and people with chronic health conditions at disproportionate risk.^2,3^ Therefore, electronic health records (EHR) with longstanding capture of patients’ medical history are uniquely positioned to answer population-based questions related to healthcare resource utilization, economic impact, and pharmaceutical intervention associated with COVID.

The Clinical Practice Research Datalink (CPRD) is a longitudinal and anonymized EHR database of primary health care interactions in England.^4^ Primary care is the cornerstone of healthcare in England, with over 98% of the population registered with a general practitioner (GP). From August 2020 through March 2022, the National Health Service (NHS) mandated that all polymerase chain reaction (PCR) tests for SARS-CoV-2, regardless of result, be reported to patients’ GP.^5^ Similarly, COVID vaccines administered at any location in the country were also mandated to be reported to patient’s GP.^6^ Thus, during this period, CPRD can be considered a closed network of confirmed COVID cases for persons in the network, with accurate information on COVID vaccination status.

While the CPRD has been used in over 3,000 peer-reviewed publications, it is not known whether interruptions in healthcare, particularly in-person primary care visits, during the first years of the coronavirus pandemic affected the previously described characteristics of the CPRD population. The objective of this study was to create case definitions for COVID diagnoses, COVID vaccination, and three separate definitions of high risk of severe COVID. Second, we aimed to evaluate these definitions in a cohort of persons with COVID in the CPRD to assess whether their sociodemographic and clinical characteristics reflected COVID epidemiology in England. The methodology developed in the study can be leveraged in future COVID research in CPRD data.

## METHODS

### Study Setting and Population

CPRD Aurum contains data which is routinely collected from primary care practices that use the EMIS Web® digital clinical system that includes electronic patient records.^7^ The May 2022 release of CPRD Aurum contained data from approximately 24% of persons in England.^8,9^ Data captured include age, sex, body weight, medical diagnoses, referrals to specialists and/or secondary care, prescriptions issued in primary care, laboratory tests, vaccinations administered, smoking and alcohol consumption status, and all other types of care delivered as part of routine primary care practice.

CPRD Aurum was linked to Hospital Episode Statistics Admitted Patient Care (HES APC) records, using deterministic patient-level linkage with an 8-stage algorithm carried out by NHS Digital. Over 99% of practices contributing to CPRD Aurum participate in HES linkage. The HES APC dataset includes patient demographics, date and method of hospital admission and discharge, diagnoses, specialty care, and procedures. HES APC data from April 1997 to March 2021 was available in this study.

### Inclusion and Exclusion Criteria

We included persons of any age diagnosed with COVID (described below) from 1 August 2020 through 31 January 2022. First, we required records to be of acceptable research quality, as defined by CPRD. Second, we required people to be continuously registered with their GP practice for at least 365 days prior to COVID diagnosis, to establish pre-COVID health history. Third, we required persons to be HES APC linkage eligible to ensure the exclusion of patients where confirmed hospital admission status (via HES APC, during the time period available) could not be known. Fourth, we excluded persons who were admitted to the hospital on or before their primary care recorded date of COVID diagnosis. Due to mandatory reporting guidelines, the GP’s date of notification from the hospital may be delayed from the date of true test collection and therefore may not accurately reflect date of diagnosis. Lastly, we excluded persons with a registration end date, practice last collection date or death date that was prior to their COVID diagnosis.

### COVID Case Definition

With each monthly data release, CPRD publishes feasibility counts for SARS-CoV-2 related codes in CPRD primary care data with corresponding code lists.^8,10^ The code types include vaccination, tests (including PCR, antibody and antigen tests), diagnosis, advice, possible cases, and post-COVID clinic referral codes. Three reviewers (KA, QM, AS) independently screened the CPRD code list to determine which of the codes represented a confirmed and current infection. Discrepancies were adjudicated by a fourth reviewer (LM), and the final definition for the COVID case definition was reviewed by UK and non-UK clinicians **(eTable 1)**. We defined a current and confirmed COVID episode as a diagnosis code, positive PCR, or antigen test. We did not include COVID vaccination, antibody tests, possible cases, exposure to COVID or post-COVID clinic referral codes in the COVID case definition.

**TABLE 1:** Characteristics of COVID Cases in CPRD Aurum, 1 August 2020 – 31 January 2022.

|  | Primary Care COVID Cases<br>(n = 2,257,907) | Hospitalized COVID Cases<br>(n = 13,165) |
| --- | --- | --- |
| Age at COVID diagnosis, years |  |  |
| 1-4 | 40,658 (2%) | 15 (< 1%) |
| 5-11 | 260,186 (11%) | 21 (< 1%) |
| 12-17 | 263,800 (11%) | 24 (< 1%) |
| 18-49 | 1,161,843 (52%) | 3,127 (24%) |
| 50-64 | 379,528 (17%) | 4,844 (37%) |
| 65-74 | 92,573 (4%) | 2,386 (18%) |
| 75-84 | 40,481 (2%) | 1,690 (13%) |
| 85+ | 18,838 (1%) | 1,058 (8%) |
| Sex |  |  |
| Male | 1,046,275 (46%) | 7,537 (57%) |
| Female | 1,211,592 (54%) | 5,628 (43%) |
| Unknown | 40 (< 1%) | 0 |
| General Practitioner Practice Region |  |  |
| North East | 83,834 (4%) | 467 (4%) |
| North West | 471,195 (21%) | 2,886 (22%) |
| Yorkshire and The Humber | 71,429 (3%) | 340 (3%) |
| East Midlands | 49,651 (2%) | 214 (2%) |
| West Midlands | 377,125 (17%) | 2,065 (16%) |
| East of England | 95,139 (4%) | 414 (3%) |
| London | 424,908 (19%) | 2,984 (23%) |
| South East | 448,536 (20%) | 2,682 (20%) |
| South West | 235,785 (10%) | 1,113 (8%) |
| Unknown | 305 (< 1%) | 0 |

Hospitalizations for COVID were defined as persons admitted with a primary diagnosis of COVID (ICD-10 U07.1 “COVID-19”) within 12 weeks of the initial diagnosis recorded in primary care. Non-hospitalized COVID cases were defined as the subset of persons for whom secondary care data were available but had no record of hospital admission. Primary care COVID cases were defined as persons who were not admitted to the hospital within 12 weeks of diagnosis, and those after April 1, 2021 for which hospital admission data were not available.

### COVID Vaccination Definition

The first COVID vaccine dose in England was administered on 8 December 2020, with the initially limited supply prioritized for groups as outlined by the Joint Committee for Vaccinations and Immunisations (JCVI).^11^ In England, COVID vaccines produced by Pfizer-BioNTech and Moderna have been available since December 2020. In April 2021, JCVI announced that persons who received a first dose of an AstraZeneca vaccine would receive a second dose of the same brand but persons who had not yet received a vaccine dose would be “preferentially offered an alternative”.^12^ This study did not consider Novavax vaccinations, as it was approved for use after our index date, or the Janssen or Valneva vaccines, which have not been used in England as of February 2023.^13^

In Phase 1 (December 2020 - March 2021), people aged 50 and older, as well as front-line health and social care workers, clinically extremely vulnerable persons and persons aged 16 and older with underlying health conditions were eligible to receive two doses. In Phase 2 (beginning April 2021), access broadened for persons aged 18-49 to receive two doses. Additionally, immunocompromised persons who received two doses in phase 1 were recommended to receive a third dose at least eight weeks after their second dose in order to complete their primary series. Phase 3 included a single dose for 16–17-year-olds in August 2021. In September 2021, 12–15-year-olds could receive a single dose and a booster dose for adults was recommended at least 6 months after the completion of a primary series. In November 2021, 12–17-year-olds could receive a second dose, and the recommendation for booster dose was shortened to at least 3 months after the completion of a primary series.^11^

We used a combination of product codes, which specify brand and dose of vaccine, as well as medical codes which indicated administration of a non-specific COVID vaccine **(eTable 2)**. The code lists were derived from the published set by CPRD.^10^ While patients may have multiple vaccination codes in their record on a given day, such as a code to indicate both the administration as well as a separate code for the specific product given, persons were not counted as receiving as more than one vaccine dose per day.

**TABLE 2:** Vaccination Status at COVID Diagnosis in CPRD Aurum, 1 August 2020 – 31 January 2022.

|  | Primary Care COVID Cases<br>(n = 2,257,907) | Hospitalized COVID Cases<br>(n = 13,165) |
| --- | --- | --- |
| Non-immunocompromised | 2,193,241 (100%) | 12,984 (100%) |
| Unvaccinated | 1,170,509 (53%) | 12,744 (98%) |
| 1 COVID vaccine dose | 173,853 (8%) | 240 (2%) |
| 2 COVID vaccine doses | 598,367 (27%) | 0 |
| First booster | 250,512 (12%) | 0 |
| Immunocompromised | 64,666 (100%) | 181 (100%) |
| Unvaccinated | 1,306 (2%) | 102 (56%) |
| 1 COVID vaccine dose | 6,033 (9%) | 79 (44%) |
| 2 COVID vaccine doses | 33,953 (53%) | 0 |
| 3 COVID vaccine doses | 23,108 (36%) | 0 |
| First booster | 266 (< 1%) | 0 |

Vaccination status at COVID diagnosis date was reported, regardless of brand, and separately for immunocompromised versus non-immunocompromised persons. Persons were considered unvaccinated if there was no record of COVID vaccination, or for up to 13 days after first dose to account for time needed to build immunity following immunisation. Dose 1 was defined starting 14 days after the record of the first COVID vaccine administration until 13 days after the receipt of second dose, or until end of follow-up. Dose 2 was defined starting 14 days after the record of the second COVID vaccine administration and administered at least 21 days after the first dose. Persons were considered to have had 2 doses until 13 days before their next vaccine. For non-immunocompromised persons, the primary vaccination series was complete 14 days after second dose. For immunocompromised persons only, dose 3 was part of their primary series at least 21 days after their second dose. First booster doses (winter 2021) were defined 21 or more days after the receipt of the last dose in primary series. For all vaccination definitions, no upper limit on time between doses was applied.

### COVID Risk Status

We considered three separate definitions of COVID risk status, each of which are set forth by different groups and capture non-intersecting populations. First, the NHS highest risk group, which was a list of conditions set forth by an advisory group commissioned by England’s Deputy Chief Medical Officer to identify persons at the very highest risk of COVID hospital admission and death.^14^ Second, eligibility for the PANORAMIC study, which began in December 2021 and is a platform randomized trial of antiviral therapeutic agents.^15^ The persons who qualify for antiviral treatment in the trial are those at a higher risk of hospital admission and death. Third, UK Health Security Agency (UKHSA) clinical risk groups as outlined in “The Green Book” chapter 14a, which is COVID vaccination prioritization from the JCVI.^16^ For each of these three risk definitions, we operationalized the clinical conditions lists into SNOMED and ICD-10 code lists. Where available, we used published code lists.^17–23^ For concepts not found in the literature, we developed wildcard-based search terms in the CPRD code browser **(eTables 3-5)**. At least one practicing clinician from the UK, who were independent and external to Pfizer, reviewed each of the search term strategies, as well as ensuing code lists.

**Table 3:** Clinical Characteristics of Primary Care COVID Cases in CPRD Aurum, by High-Risk Definitions.

|  | All<br>(n = 2,257,907) | Meeting NHS Highest<br>Risk Conditions<br>(n = 249,972) | Meeting PANORAMIC<br>Criteria<br>(n = 691,593) | Meeting<br>UKHSA<br>Clinical Risk<br>(n = 225,051) |
| --- | --- | --- | --- | --- |
| Primary care cases meeting definition | -- | 11% | 31% | 10% |
| Smoking status |  |  |  |  |
| Current smoker | 298,735 (13%) | 44,365 (17%) | 121,019 (18%) | 46,436 (21%) |
| Former smoker | 285,722 (13%) | 67,637 (26%) | 168,343 (24%) | 63,124 (28%) |
| Never smoked | 580,239 (26%) | 87,217 (34%) | 228,579 (33%) | 77,157 (34%) |
| Unknown | 1,093,211 (48%) | 60,753 (23%) | 173,652 (25%) | 38,334 (17%) |
| Body mass index (kg/m<sup>2</sup>) |  |  |  |  |
| Mean (standard deviation) | 27.6 (7.1) | 29.1 (6.9) | 29.5 (6.7) | 29.5 (7.5) |
| Underweight (< 18.5) | 45,194 (2%) | 3,720 (1%) | 5,305 (1%) | 5,926 (3%) |
| Not overweight (18.5-24.9) | 226,272 (10%) | 35,766 (14%) | 81,363 (12%) | 36,716 (16%) |
| Overweight (25.0-29.9) | 204,360 (9%) | 45,261 (17%) | 113,863 (16%) | 43,758 (19%) |
| Obese (≥30.0) | 219,253 (10%) | 53,814 (21%) | 138,035 (20%) | 62,494 (28%) |
| Unknown | 1,562,828 (69%) | 121,411 (47%) | 353,027 (51%) | 76,157 (34%) |
| Charlson Comorbidity Index |  |  |  |  |
| Mean (standard deviation) | 0.2 (0.6) | 0.7 (1.3) | 0.5 (1.0) | 1.0 (1.3) |
| 0 | 2,003,257 (89%) | 165,823 (64%) | 487,550 (71%) | 102,790 (46%) |
| 1 or 2 | 221,370 (10%) | 74,805 (29%) | 174,287 (25%) | 99,830 (44%) |
| 3 or 4 | 24,554 (1%) | 12,659 (5%) | 21,574 (3%) | 15,961 (7%) |
| 5+ | 8,726 (< 1%) | 6,685 (3%) | 8,182 (1%) | 6,470 (3%) |

### Comparison to Publicly Available Data

We compared our results to estimates published on the UK Government’s Coronavirus dashboard, and restricted results to those specific to England, as the CPRD Aurum population that is HES-linkage eligible contains data from England only.^24,25^ The data in this manuscript reflect figures from the dashboard as accessed on 31 January 2023.

Where possible, we restricted the dashboard to cases from 1 August 2020 through 31 January 2022 to reflect the study period, although age, sex and region-specific estimates were reported “since the start of the pandemic”.^24,25^ For whole country (non-COVID related) comparisons, we used the Office of National Statistics’ 2021 Census.^9^

### Statistical Analyses

All results are presented separately for the cohorts of persons with primary care versus hospitalized COVID cases. Continuous variables are presented as means (standard deviations) or medians (interquartile ranges) and categorical variables as counts (percentages). Missing data for sex, region, smoking status and BMI is shown in tables. The absence of codes for comorbidities was assumed to be the absence of the comorbidity rather than missing data. Standardized mean differences (SMD) were used to compare groups, with SMD > 10% indicating significant difference. As per CPRD privacy rules, any cell with 10 persons or fewer (but not zero) was suppressed and other cells related to the small cell count were redacted to ensure no back-calculation could populate the count.

CPRD obtains ethical research approval annually from the UK’s Health Research Authority Research Ethics Committee to accumulate and distribute patient data. This study is based in part on data from the Clinical Practice Research Datalink obtained under license from the UK Medicines and Healthcare products Regulatory Agency. The data is provided by patients and collected by the NHS as part of their care and support. Office for National Statistics (ONS) and Hospital Episode Statistics (HES) data, copyright © (2021), re-used with the permission of The Health & Social Care Information Center. All rights reserved. The interpretation and conclusions contained in this study are those of the authors alone. Data management and analyses for this study used SAS, version 9.4 (SAS Institute, Cary, North Carolina).

There were no directly involved patients or public involvement in this study.

## RESULTS

From 1 August 2020 through 31 January 2022, the UK Government’s Coronavirus dashboard reported 14,744,991 COVID cases in England.^24^ The final CPRD cohort contained 2,271,072 persons diagnosed with COVID, regardless of care setting, in England in the same time period **(eTable 6)**. The case trends followed national estimates, with a series of peaks and troughs with a large peak in winter 2021 **(eFigures 1 and 2)**. Hospital admissions increased in fall 2020 and winter 2020 **(eFigure 3)**.

**Figure 1:**
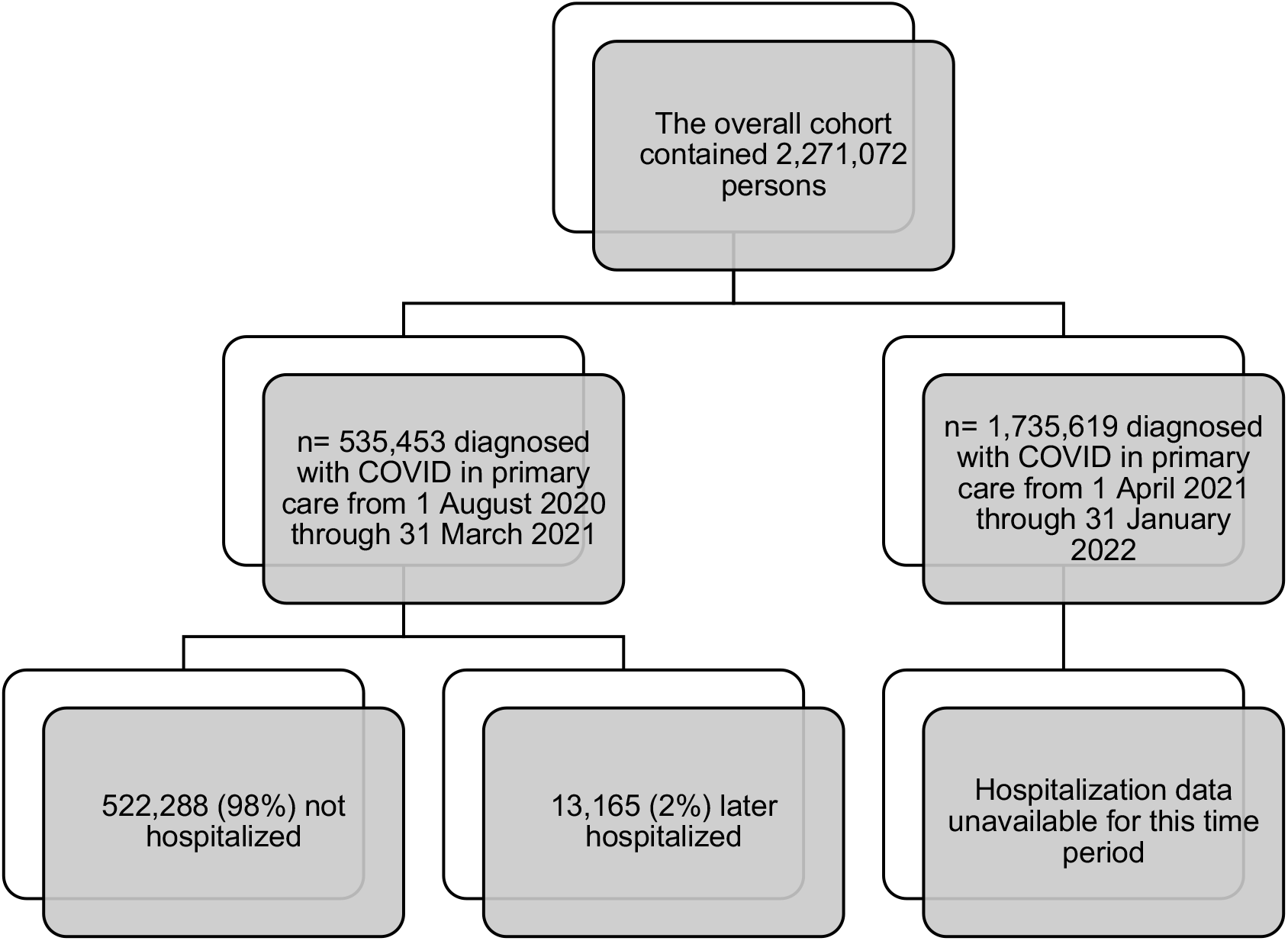
Cohort Description.

There were 2,257,907 persons with COVID cases observed in primary care, which included persons who were not hospitalized during the period of time for which hospital admission data was available (n=522,288) as well as all COVID diagnoses recorded in the period where hospital admission data ended (n=1,735,619). Separately, there were 13,165 persons (2% of COVID cases from August 2020 to March 2021) who were hospitalized within 12 weeks of initial COVID diagnosis with a primary diagnosis of COVID **(Figure 1)**.

### Characteristics of COVID Cases

For both primary care and hospitalized cases, the majority of persons with COVID were adults aged 18-64 **(Table 1)**. The age distribution of CPRD primary care cases generally followed the national case count distribution, with young and middle-aged adults representing the largest groups, as well as the largest fraction of the English population **(eTable 7)**. Older adults were underrepresented in both cohorts as compared to national population estimates. Within the primary care cohort, 6% of patients were aged 65-84 years, whereas 9% of national COVID cases occurred in this age group. Notable differences were observed in the hospitalized cohort when compared to the national estimates; 31% vs. 40% were aged 65 - 84 years, and 8% vs. 21% aged ≥ 85 years, respectively.

The sex distribution of COVID cases in the primary care cohort was comparable to national estimates, with 55% female in each **(Table 1 and eTable 7)**. There were more males (57%) than females in the hospitalized cohort; there are no national sex-specific hospital admission counts available.

The regional distributions of the CPRD primary care and hospitalized cohorts were similar to each other, but not the overall country. Both of the study cohorts had larger proportions of persons with GP practices in higher income regions of England as compared to national case counts.^26^ For example, 20% of persons in the primary care cohort and 20% in the hospitalized cohort lived in South East region of England, the region with the highest median total household wealth as reported in March 2020. This is compared to 20% of CPRD contributing practices, 15% of all English cases and 16% of the population lived in London **(eTable 8)**. Similar trends are seen with lower median total household wealth regions being proportionally underrepresented in the COVID CPRD cohort as compared to national case estimates or population distribution.

### COVID Vaccination Status

At the time of COVID diagnosis, 27% of non-immunocompromised persons and 36% of immunocompromised persons had completed a primary series of COVID vaccination **(Table 2)**. Among the non-immunocompromised hospitalized COVID cases, 98% were unvaccinated and none had completed a primary series. For immunocompromised hospitalized COVID cases, 56% were unvaccinated, 44% had received one dose, and none had completed a primary series. As of 1 December 2021, 86% of adults had ≥1 and 80% had ≥2 COVID vaccine doses (2% and 0.2% lower than official reports, respectively).

### Populations at Risk of Severe Disease

After wildcard searches, clinicians reviewed nearly 50,000 codes to set the definitions for high-risk lists. The final lists contained 12,390 codes for NHS Highest Risk, 9,132 codes for PANORAMIC trial criteria and 7,343 codes for UKHSA Clinical Risk **(attached as machine readable code list files)**. In the primary care cohort, 11% met NHS Highest Risk, 31% PANORAMIC, and 10% UKHSA Clinical Risk criteria **(Table 3)**. With each definition, primary care cohort cases at high risk were more often current smokers (ranging from 17-21% vs 13% in full cohort) or former smokers (24-28% vs 13%), overweight (16-19%, vs 9%) or obese (20-28%, vs 10%), and a larger proportion had at least 1 comorbidity in the Charlson Index (29-54%, vs 11%), where this was recorded. Primary care cases at high risk of severe disease were also older (mean age 49-55 years, vs 34 years), more often female (55-63%, vs 54%), and had more recorded vaccine doses prior to COVID diagnosis **(eTable 9)**.

Among hospitalized COVID cases, 33% met NHS Highest Risk, 84% PANORAMIC, and 41% UKHSA Clinical Risk criteria’s **(Table 4)**. The high risk hospitalized groups were similar to the overall hospitalized cohort. Nearly half of persons were current smokers (13-16%, vs 13%) or former smokers (32-39%, vs 29%), and more than half were overweight (20-23%, vs 18%) or obese (37-42%, vs 35%). Among the hospital admitted cohort, people meeting each high-risk definition were older (mean age 65-66 years, vs 60 years) **(eTable 10)**. Subgroups of patients with the NHS and UKHSA definitions had higher mean Charlson Comorbidity Indexes, and more females, than high risk patients identified with the PANORAMIC criteria or the entire hospitalized population.

**Table 4:** Clinical Characteristics of Hospitalized COVID Cases in CPRD Aurum-HES Linked Data, by High-Risk Definitions.

|  | All<br>(n = 13,165) | Meeting NHS Highest Risk<br>Conditions<br>(n = 4,333) | Meeting<br>PANORAMIC<br>Criteria<br>(n = 11,011) | Meeting<br>UKHSA<br>Clinical Risk<br>(n = 5,353) |
| --- | --- | --- | --- | --- |
| Hospitalized cases meeting definition | -- | 33% | 84% | 41% |
| Smoking status |  |  |  |  |
| Current smoker | 1,690 (13%) | 655 (15%) | 1,476 (13%) | 840 (16%) |
| Former smoker | 3,810 (29%) | 1,574 (36%) | 3,544 (32%) | 2,070 (39%) |
| Never smoked | 4,205 (32%) | 1,362 (31%) | 3,513 (32%) | 1,746 (33%) |
| Unknown | 3,460 (26%) | 742 (17%) | 2,478 (23%) | 697 (13%) |
| Body mass index (kg/m<sup>2</sup>) |  |  |  |  |
| Mean (standard deviation) | 31.9 (7.5) | 31.4 (7.5) | 31.7 (7.4) | 31.4 (7.7) |
| Underweight (< 18.5) | 96 (1%) | 51 (1%) | 90 (1%) | 70 (1%) |
| Not overweight (18.5-24.9) | 1,162 (9%) | 509 (12%) | 1,060 (10%) | 729 (14%) |
| Overweight (25.0-29.9) | 2,390 (18%) | 949 (22%) | 2,161 (20%) | 1,239 (23%) |
| Obese (≥30.0) | 4,586 (35%) | 1,662 (38%) | 4,053 (37%) | 2,231 (42%) |
| Unknown | 4,931 (37%) | 1,162 (27%) | 3,647 (33%) | 1,084 (20%) |
| Charlson Comorbidity Index |  |  |  |  |
| Mean (standard deviation) | 1.0 (1.6) | 1.9 (2.1) | 1.2 (1.7) | 2.0 (2.0) |
| 0 | 7,128 (54%) | 1,283 (30%) | 5,158 (47%) | 894 (17%) |
| 1 or 2 | 4,241 (32%) | 1,833 (43%) | 4,094 (37%) | 2,927 (55%) |
| 3 or 4 | 1,161 (9%) | 711 (16%) | 1,129 (10%) | 963 (18%) |
| 5+ | 635 (5%) | 506 (12%) | 630 (6%) | 569 (11%) |

## DISCUSSION

### Key Results

In this work, we defined and benchmark results from three key variables related to COVID research using CPRD: index COVID diagnoses, COVID vaccinations, and persons at high risk of severe disease.

We identified 2,271,072 COVID cases in CPRD Aurum between 1 August 2020 through 31 January 2022. Younger age and lower socioeconomic deprivation have been consistently associated with reductions in COVID incidence and severity.^27,28^ These factors may explain why this CPRD cohort, which proportionally underrepresented persons age 65 and older and overrepresented persons living in the regions with higher median total household wealth, captured 15% of COVID cases in a database that covers 24% of persons in England. Future work using CPRD for COVID research will need to consider these limitations of under ascertainment of cases. Moving beyond this study’s time period, the transition to at home testing, as well as the end of free PCR testing for the general public on April 1, 2022, will need to be additionally considered.

COVID vaccination events were well captured in the CPRD. This stands in stark contrast to most administrative claims and EHR databases in the United States, where less than 50% of COVID vaccines are recorded in comparison to estimates provided by the Centers for Disease Control and Prevention.^29,30^ England’s national healthcare system, as well as the NHS data infrastructure to facilitate capture of COVID-related events and longstanding structure of general practitioners as the central node in a person’s healthcare coordination, enabled the high coverage of COVID vaccination records. Researchers can be more confident with CPRD data that the absence of a vaccination record indicates unvaccinated status than they otherwise would be with most other real-world datasets, which is a critical consideration for studies related to COVID disease burden, vaccination, or treatments.

The proportion of persons who had completed primary series vaccination prior to infection was low among primary care cases. Notably, among hospitalized cases, no patients had completed a primary COVID vaccination series. These findings may be explained by several factors. First, the COVID vaccine was first made available in England on 8 December 2020, and initially second doses were given up to 12 weeks later to maximize limited supply for as many people as possible. Therefore, the calendar period under study allowed for many persons to have had a COVID diagnosis in periods at which “full vaccination” was not achievable. Second, it is possible that one vaccine offered protection against severe illness.^31–33^

We operationalized a total of over 28,000 codes, from an initial set of nearly 50,000, for three definitions of persons at risk of severe disease. We have made the search terms available, for reproducibility, as well as the resulting code lists, for other research groups to implement in their work. To our knowledge, we offer the first publication of code lists for each of these high-risk definitions, which can now be readily used in datasets that contain CPRD medical and product, ICD-10 and OPCS Classification of Interventions and Procedures codes.

While there are similarities between the three definitions, differences do exist. In the example of renal disease, NHS Highest Risk defines as chronic kidney disease stage 4 or 5, PANORAMIC trial criteria stipulate stage 2 or 3, and UKHSA Clinical Risk are for stages 3-5. PANORAMIC trial eligibility capture persons with mild renal disease, as some antiviral treatments are not approved for use in persons with severe renal disease. However, UKHSA Clinical Risk prioritized vaccination access for persons at highest risk of disease, which would include persons with more advanced renal disease. Notably, persons who have renal disease as defined by PANORAMIC trial criteria by definition do not have renal disease by NHS definition, and people in each of these may (or may not) meet UKHSA prioritization. The choice of which high risk definition to implement in future studies will need to be guided by the study population and research question.

In the primary care cases, persons at high risk were older, more often smokers, had larger body sizes, and higher Charlson Comorbidity Indices from the overall group of primary care cases. Among hospitalized cases, the high-risk groups were similar to the entire hospitalized group. These findings are consistent with existing understanding of high-risk definitions, and perhaps provide reassurance that the code lists measure the purported phenomenon.

The study periods in this report represent the most recent data available from CPRD as of 3 February 2023. During the Autumn of 2022, the Aurum database experienced data quality issues related to the EMIS data flows from legacy systems, and no primary care data have been made available to researchers since the May 2022 release (data through March 2022, with some early view of April 2022). Separately, HES secondary care data have not been updated since March 2021, as NHS Digital has undergone a change in the way they process and link data. COVID remains a serious disease for some people, and it is certain that some of the 1.7 million persons diagnosed with COVID after April 1, 2021 would have been later admitted for COVID, but we do not have the hospital admission data to distinguish them from those whose cases were managed entirely in the community setting. Throughout, we have used the term “primary care records” as the combined groups of those known to be non-hospitalized (cases where HES data were available, but the person was not hospitalized), as well as those who we have GP encounters for but unknown eventual hospital admission status. It is difficult to approximate the number of hospital admissions that would be expected with full data availability. Carrying forward the 2% hospital admission incidence seen in the early pandemic period may not be appropriate, given 2021 introduced periods of increased (delta variant) and decreased (omicron variant) risk of hospital admission, as well the uptake of COVID vaccinations and antiviral treatments. Finally, the population structure of this cohort outlined in this work further challenge the direct application of national estimates to CPRD cohorts.

The present study does not capture persons not under GP care such as prisoners, some residential homes, and persons without a place of residence. Additionally, CPRD Aurum, when linked with HES data, reduces the population to persons registered at eligible GP practices in England and therefore may not represent persons in other countries in the UK, or countries outside the UK. This study does not include persons who presented directly to hospital without any prior GP interaction. In particular, persons with more severe disease such as older adults may require immediate hospital admission before seeking primary care, which could explain some of the gaps in representation we have reported. Given that CPRD is a primary care database, and the limited time period of hospital data availability, we decided to design our study as an initial cohort of persons with primary care records of COVID. Studies looking for complete capture of all hospitalized COVID patients might consider other data sources.

In conclusion, we present a cohort of over 2 million COVID cases in linked CPRD-HES data, using published definitions for COVID cases, vaccinations, and each of three UK-specific definitions of persons at high risk of severe disease. CPRD primary care data is a robust real-world data source and can be used for some COVID research questions; however, limitations of the data availability should be carefully considered.

## Data Availability

Electronic health records are considered sensitive data in the UK by the Data Protection Act and cannot be shared. The primary care data can be requested via application to the Clinical Practice Research Datalink, with secondary care data and mortality data through linkage upon application. Information is available from https://www.cprd.com/research-applications.

## Conflict of Interest Statement

Dr Andersen, Dr McGrath, Ms Reimbaeva, Dr Mendes, Dr Nguyen, Dr Tsang, Ms Malhotra and Dr Yang are employees of Pfizer Inc or Pfizer Ltd and may hold Pfizer stock or stock options. Dr Rai and Mr Tritton are employees of Adelphi Real World which has received consulting fees from Pfizer Inc.

The authors gratefully acknowledge Bethany Backhouse, Poppy Payne, Elke Rottier and Robert Wood from Adelphi Real World (Bollington, UK), Tamuno Alfred, Darren Kailung Jeng, Tendai Mugwagwa, Qiao Mu and Chern Chuan Soo from Pfizer Inc. (New York, United States) Agnieszka Gajewska, Tomasz Mikołajczyk and Ewa Śleszyńska-Dopiera from Quanticate (Warsaw, Poland), and Andy Surinach from Genesis Research (Hoboken, United States).

## Transparency Statement

Dr Andersen affirms that this manuscript is an honest, accurate, and transparent account of the study being reported; that no important aspects of the study have been omitted; and that any discrepancies from the study as planned (and, if relevant, registered) have been explained.

This study reports pre-specified analyses according to a registered protocol with CPRD (CPRD study number 22_002062) and Pfizer documented statistical analysis plan (study number KIMS 3106).

